# Lung function changes of divers after single deep heliox diving

**DOI:** 10.1101/2021.03.10.21253337

**Authors:** Xiao-Chen Bao, Yi-Qun Fang, Tao Yang, Yong-jun Sun, Jun Ma, Ji Xu, Nan Wang, Fang-Fang Wang

## Abstract

**Objectives:** This study detects the changes in pulmonary function of divers after 80m, 100 m, and 120 m helium-oxygen (heliox) dive. Methods: A total of 26 divers participated in the experiment, of which 15 divers performed the 80m dive, 5 divers performed the 100m dive, and 6 divers performed the 120m dive. The exposure phases included breathing heliox or air in water and O_2_ in the hyperbaric chamber. Pulmonary function (forced flow-volume) was measured twice before diving, within 30 minutes after diving, and 24 hours after diving. The parameters examined were forced vital capacity (FVC), forced expired volume in 1 second (FEV1), forced expired flow from 25% to 75% volume expired (FEF25-75%), 25-75 percent maximum expiratory flow as compared with vital capacity (MEF 25-75%) and peak expiratory flow (PEF). Results: FEV1/FVC and MEF25% markedly decreased (p = 0.0395, *p* = 0.0496) within 30min after the 80m dive, but returned to base values at 24h after the dive. Other indicators showed a downward trend within 30min after 80m heliox diving (no statistical difference). Interestingly, FEV1, FEV1/FVC, PEF, MEF decreased after 100m heliox dives, but there was no statistical difference. However, in the 120m heliox dive, FEV1/FVC and MEF75% significantly decreased again after diving (*p* = 0.0098, *p* = 0.0073). The relatively small number and more proficient diving skills of divers in 100m and 120m diving may be responsible for the inconsistent results. But when the diving depth reached 120m, results again showed a significant statistical change. Conclusion: Single deep heliox diving can cause temporary expiratory and small airway dysfunction, which can be recovered at 24h after diving.

## Introduction

Diving is a special operation with high risk. Diving environment, such as elevated ambient pressure, altered gas characteristics, changes in cardiovascular stress after immersion in water and other factors, all have an impact on the lung1. It has been found that long-term deep diving can cause lung small airway disease and decrease lung function of commercial divers^2 3^, but accelerated decline in lung function were not found in military and recreational divers ^4-6^. For clinically susceptible individuals, a single dive can change pulmonary function^7 8^. Effects of submergence, static lung loading, as well as effects of respiratory heat and water loss, may contribute to the adverse effects of a single wet scuba dive on pulmonary function^8 9^.

Helium is less soluble and more diffusive than nitrogen. A mixture of helium and oxygen, termed heliox, are used as breathing medium in deep dive to avoid the narcotic effects of nitrogen under pressure ^10^. The lower density of heliox than nitrogen and oxygen mixture (air, or airox) seemed to has beneficial effects in diving. Breathing heliox leads to a reduction in resistance to flow within the airways, and consequently to a decrease in the work of breathing ^11 12^. However, increasing respiratory heat loss in heliox diving and long decompression time under water have negative effects on human respiratory system. There are few studies of the effects of heliox diving on human pulmonary function. Marinovic reported that trimix dive (55 to 80 m) was associated with accumulation of extravascular lung water, and diminished left ventricular contractility ^13^. Decreasing transfer factor for carbon monoxide was observed after eight saturation dives to pressures of 3.1-4.6 MPa^14^. However, information of the effect of a single deep heliox diving (more than 80m sea water) on human pulmonary function is unknown.

The aim of this study was to detect the changes in lung function of divers after a single deep heliox dive.

## Methods

### Subjects

Twenty-six male healthy divers were recruited for this investigation. Fifteen of them performed the 80 m heliox dive, five divers performed 100 m, and six divers performed the 120 m heliox dive. Participant characteristics are summarized in Table 1. All experimental procedures in the study were completed in accordance with the Declaration of Helsinki and approved by Ethical Committees of the Naval Medical Center. All subjects gave informed written consent. Health status and previous diving experience was self-reported.

**Table 1.** Characteristics of divers participating in heliox diving at different depths

|  | 80m | 100m | 120m |
| --- | --- | --- | --- |
| Age (years) | 24.07±4.57 [18-32] | 29.33±2.80 [26-33] | 27.5±2.74 [24-31] |
| Body mass (kg) | 71.33±9.60 [60-88] | 72.83±5.98 [69-85] | 66.83±4.22 [60-72] |
| Height (cm) | 174.33±4.08 [170-183] | 173.33±3.20 [168-178] | 172.01±1.73 [170-175] |
| Experience(years) | 5.20±2.86 [2-12] | 10.83±3.06 [7-15] | 13.00±2.19 [10-16] |

Data are expressed as means ± SD.

### Dive protocol

The diver, breathing air to heliox (He: O_2_, 82:18), with gradual immersion to 80, 100 and 120 m of seawater (with the descending velocity of 15-20 m/min). The bottom time was 15min, then divers returned to the first stop station with ascending velocity of 8 m/min. After converting breathing gas from heliox to air at the first stop station, divers descend to 12m following the decompression table (Fig 1). After completing the decompression stop at 12m seawater, divers returned to the surface within 6 minutes, and then recompressed to 15m in hyperbaric chamber and breathed oxygen. The interval time between surface and recompression was controlled in 5 min. The recompression procedure is listed in Fig 1.

**Fig. 1.**
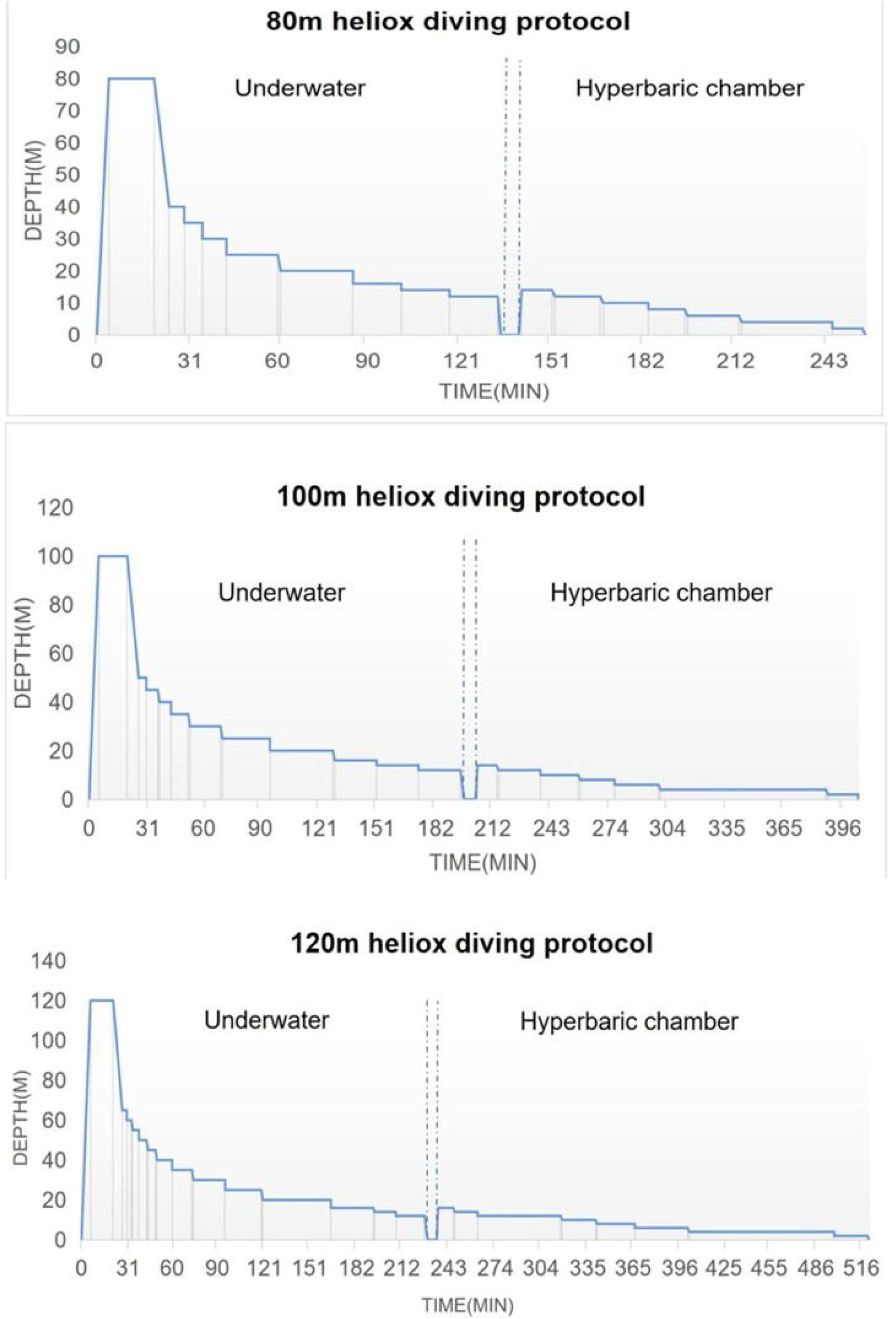
80-120m Heliox Diving Decompression procedure.

### Pulmonary function test

For all dive series, pulmonary function was measured during the week before the dives (baseline), within 30 minutes after divers surfaced, and 24hours after the dive series was complete. Forced vital capacity maneuvers were recorded using a pulmonary function machine (COSMED, Inc., Roma, Italia). The average of parameters from three flow-volume loops reproducible according to American Thoracic Society standards^15^ was used at each time point, and was compared to a baseline average of three reproducible values which were obtained in the morning before dive.

### Statistical analysis

Statistical analyses were performed with the SPSS statistical package, version 11.5 (SPSS, Inc., Chicago, IL, USA). Results are presented as means ± standard error of mean (SEM) and means ± standard deviation (SD). The t test (2-tailed) was performed based on the normality of the distribution as checked by the Shapiro-Wilk test. A *P* value of < 0.05 was considered statistically significant.

## Results

### Effect of 80m heliox diving on diver’s pulmonary function

The mean values of pulmonary function parameters are listed in Fig 1. When compared with baseline values, FEV1/FVC% significantly decreased (87.14 ± 7.69 vs 89.19 ± 8.40, *P* = 0.0395) within 30minutes after diving, and returned to the baseline at 24 hours after diving (Fig. 2). The same trend was seen in MEF25% (2.35 ± 0.67 vs 2.57 ± 0.82, *p* = 0.0496). FEV1. FEF25-75% and MEF 50/75% slightly decreased immediately after diving when compared with baseline values, but had no statistical difference(Fig. 2). Other indicators did not change significantly before and after the dive.

**Fig. 2.**
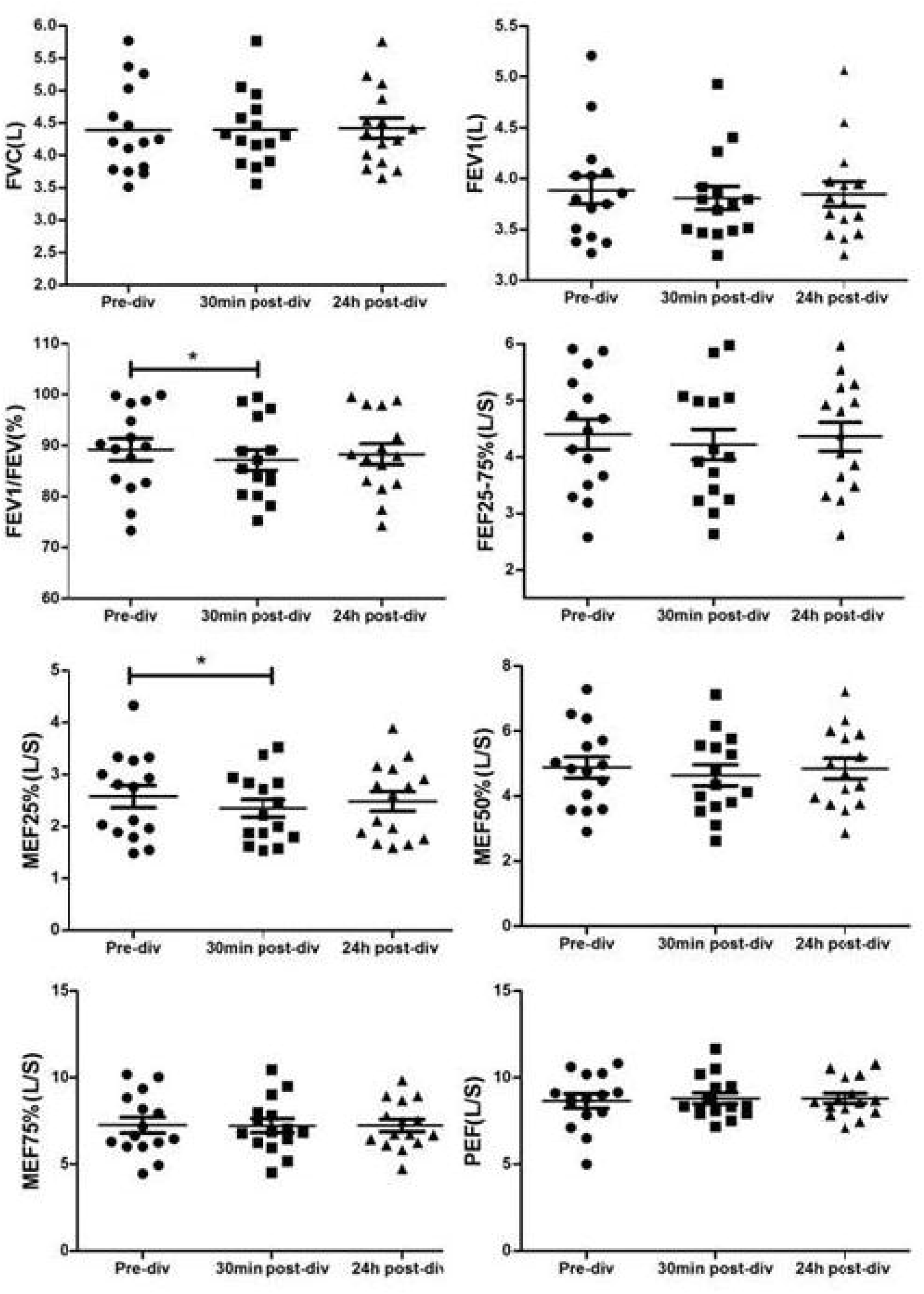
The changes of pulmonary function parameters within 30min and 24h after 80m heliox diving. \**P* < 0.05 compared to pre-dive group. Data are expressed as means ± SEM (n = 15).

### Effect of 100m heliox diving on diver’s pulmonary function

Figure 3 showed that after a 100m heliox dive, compared with the basic value before the dive, all pulmonary function parameters of divers showed a downward trend, but not significantly (*P* > 0.05). FVC, FEV1 and FEV1/FVC returned to the basic values by 24 h after diving, while PEF, FEF 25/75% and MEF 25/50/75% decreased further (Fig 3).

**Fig. 3.**
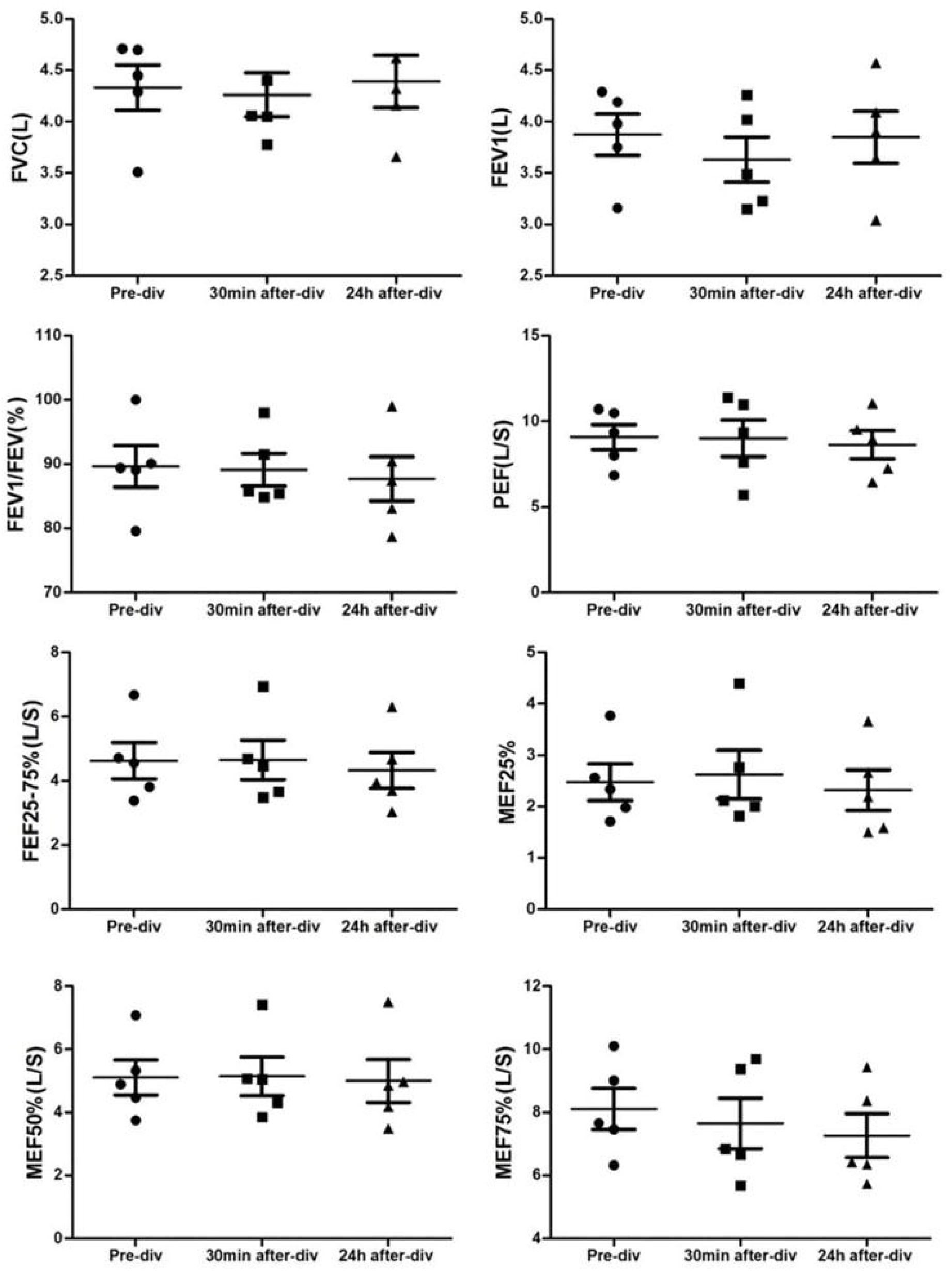
The changes of pulmonary function parameters within 30min and 24h after 100m heliox diving. Data are expressed as means ± SEM (n = 5).

### Effect of 120m heliox diving on diver’s pulmonary function

Results of 120m heliox dive are consistent with that of 80m dive. As Fig 4 showed, when compared with data before diving, FEV1/FVC% markedly decreased (85.65 ± 3.12 vs 90.36 ± 4.26, *P* = 0.0098) within 30minutes after diving, and returned to the baseline at 24 hours after diving. The changes of MEF75% presented the same trend. MEF75% decreased significantly within 30 minutes after diving when compared with values before diving (7.22 ± 1.13 vs 8.05 ± 1.17, *P* = 0.0073), but returned to baseline at 24h after diving. Other indicators did not change significantly before and after the dive.

**Fig. 4.**
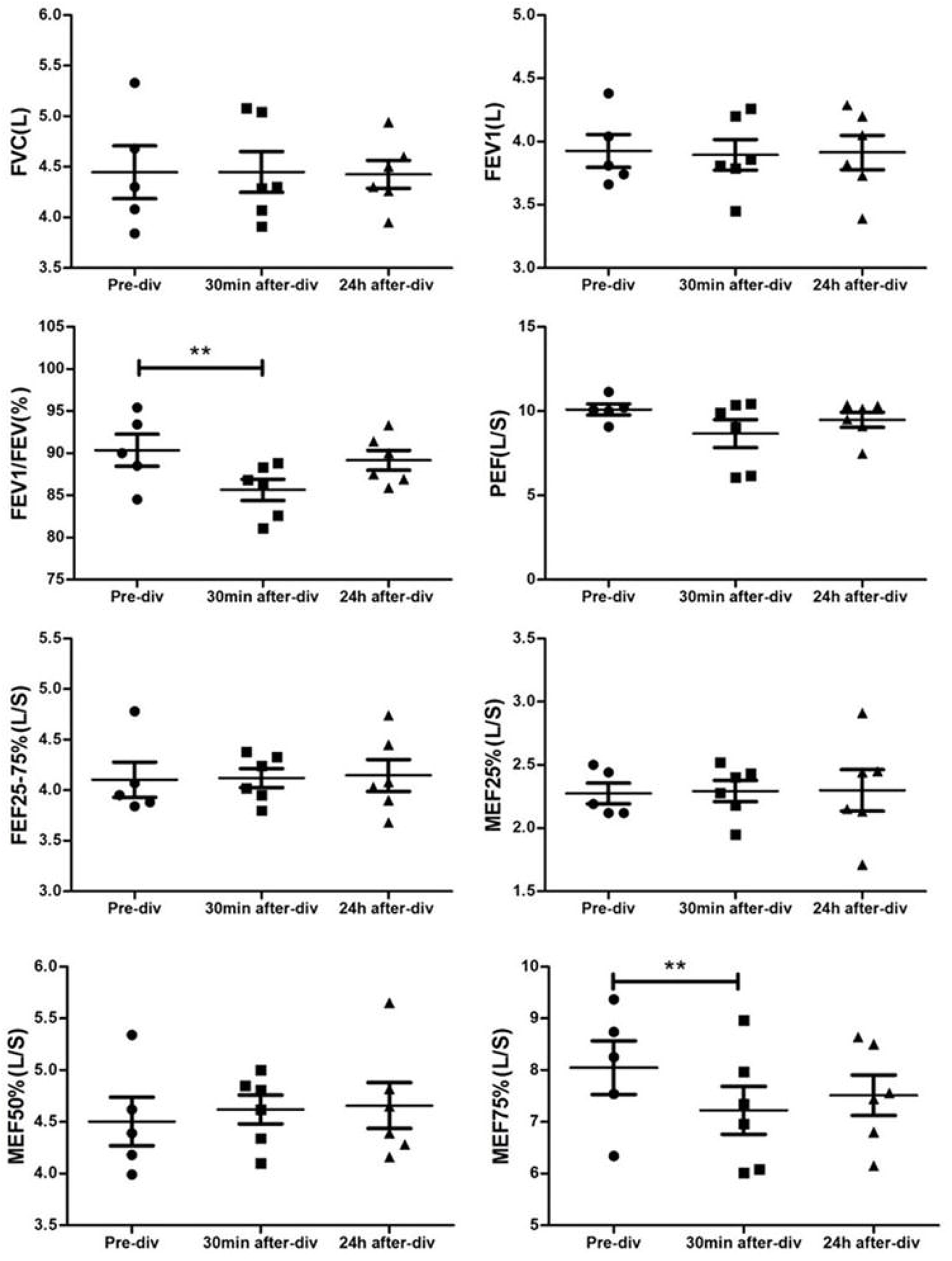
The changes of pulmonary function parameters within 30min and 24h after 120m heliox diving. \*\**P* < 0.01 compared to pre-dive group. Data are expressed as means ± SEM (n = 6).

## Discussion

This study is the first time to investigate the effects of single deep heliox diving on pulmonary function of divers. Statistical analysis showed that 80 m helium-oxygen diving can significantly decreased the FEV1/FVC% and MEF25% of divers, while these changes can return to the base value 24 hours after diving. Results of 120m heliox dive are consistent with that of 80m dive. FEV1/FVC% and MEF75% decreased markedly within 30min and returned to base value at 24h after diving.

However, results of 100m dive showed that pulmonary parameters presented a decrease tendency within 30 minutes after diving, but had no statistical difference. There are two possible reasons for these inconsistent results: 1. Large interindividual variability existed in divers’ lung function, and statistical differences can be obtained when the sample size is large (such as 15 cases in 80 m heliox dive). Considering the safety of diving and the proficiency of divers, the number of divers participating in 100m and 120m diving is small (n=5-6). Therefore, the results only showed a downward trend, but had no statistical differences; 2. The divers participating in 100m and 120m diving have more diving experience and proficient diving skills. As a result, they did less ineffective breathing during diving and had less changes in lung function. But when the diving depth reached 120m, the results again showed a significant statistical change. This result further suggests that a single deep heliox dive can cause transient and significant changes to divers’ lung function.

FEV1/FVC% is the main index to judge expiratory dyspnea. The decrease of FEV1/FVC% indicates that divers have obstructive airway changes. It has been confirmed that long-term diving can reduce the FEV1/FVC ratio of divers^16^. Increased FVC is the main reason for the decrease in FEV1/FVC^17^. In a single dive, oxygen poisoning is the main factor in reducing FEV1/FVC. For example, Shykoffbe found that breathing pure oxygen at 13m can significantly reduce FEV1 and FEF 25-75%, while breathing air had no effects ^18^. Thorsen found transfer factor for carbon monoxide (TLCO) and forced midexpiratory flow rate decreased markedly after deep saturation dives. Hyperoxia contributed significantly to these changes^19^. In our study, the oxygen inhalation time in hyperbaric chamber after 80m heliox dive was 94 minutes. The oxygen inhalation time after 100m and 120m heliox dives is longer, reaching 176 and 250 minutes, respectively. Oxygen toxicity may be an important factor which affected FEV1/FVC in this study.

MEF 25-75% decrease is common in obstructive and restrictive disorders, and also in small airway diffuse lesions. The long-term effect of diving and single diving on lung function is mainly manifested in the decrease of lung diffusion function and lung compliance^2^. In recent years, studies found that long-term diving can increase divers’ FVC value, decrease FEF25-75%, and can induce obstructive ventilation dysfunction^20^. A cross-sectional sample of 180 healthy male divers and 34 healthy male controls revealed that divers had a lower mid-expiratory flow at MEF25-50% than controls. These changes were inversely related to diving years, indicating long-term effects of diving on respiratory function ^21^. Oxygen toxicity, inert gas micro bubbles produced during decompression, stress response of cardiovascular system and increase of expiratory resistance load caused by breathing high-density dry and cold gas can all lead to the changes of pulmonary diffuse function. In our study, the decreased MEF25/75% indicates that a single deep heliox dive can cause lung small airway function damage, but the decrease is transient and can be restored after 24 hours.

The possible causes of pulmonary function changes of deep helium oxygen diving are considered as follows: 1. Oxygen toxicity: As we mentioned earlier, oxygen poisoning caused by long-term oxygen inhalation is one of the factors; 2. Increased respiratory resistance: When divers inhaled oxygen in the hyperbaric chamber, they complained of dry breathing gas and high respiratory resistance, which increased the exhalation resistance load; 3. Immersion can enrich blood to the chest and diaphragm, resulting in decreased lung volume, increased airway resistance and decreased lung compliance. However, further experiments are needed to confirm the existence of these possible factors.

## Data Availability

The data used to support the findings of this study are available from the corresponding author upon request.

